# Explaining the Effective Reproduction Number of COVID-19 through Mobility and Enterprise Statistics: Evidence from the First Wave in Japan

**DOI:** 10.1101/2020.10.08.20209643

**Authors:** Yoshio Kajitani, Michinori Hatayama

## Abstract

This study uses mobility statistics combined with business census data for the eight Japanese prefectures with the highest COVID-19 infection rates to study the effect of mobility reductions on the effective reproduction number (i.e., the average number of secondary cases caused by one infected person). Mobility statistics are a relatively new data source created by compiling smartphone location data. Based on data for the first wave of infections in Japan, we found that reductions targeting the hospitality industry were more effective than restrictions on general business activities. Specifically, we found that to hold back the pandemic (that is, to reduce the effective reproduction number to one or less for all days), a 20-35% reduction in weekly mobility is required, depending on the region. A lesser goal, 80% of days with one or less observed transmission, can be achieved with a 6-30% reduction in weekly mobility. These are the results if other potential causes of spread are ignored; more careful observations and expanded data sets are needed.

## Introduction

Many countries have suffered from the COVID-19 pandemic and have experienced severe economic impacts due to the restrictions on socio-economic activities. GDP losses have been significant (e.g., -7.9% in Japan during the second quarter of 2020 [1]), and unemployment numbers are increasing. Several countries have managed to restart socio-economic activities to near pre-pandemic levels, but most have suffered a second wave of the pandemic.

The conditions of first, second, and higher-round waves can differ because individual or organizational countermeasures (e.g., masks, hand washing, antiseptic solutions, and partitioning) have advanced. However, analyzing the infection risks and degree of lockdown/voluntary restriction of socio-economic activities in the first wave, the only currently available data, is meaningful for creating better activity restriction policies.

Traffic flows or mobility habits are a representative measure of lockdown restriction levels. Therefore, many previous studies have focused on the relationship between infection levels and traffic flows to demonstrate the effects of lockdowns/voluntary restrictions. For example, focusing on traffic data obtained from 1200 automatic traffic sensors in Italy, Cratenì et al. [2] constructed a multiple regression model to explain the number of daily new positive cases of COVID-19. In their regression model, the explanatory variables on particulate matter pollutant, number of tests per day, travel time decay from outbreak, and temperatures are statistically significant, but the mobility habits 21 days before an onset is shown to be most influential. Similarly, rail-based transport accessibility [3] and traffic volumes on express ways [4] also have the capability to explain the number of infections.

Person-based mobility statistics, which have recently become available through smartphone devices, are a powerful tool for understanding regional overviews of socio-economic activities. For example, Google Mobility Report [5] provides population statistics for retail and recreation, grocery and pharmacy, parks, transit stations, workplaces, and residential areas all over the world. Engle et al. [6] used GPS locational data for 94,116 observations in 3,142 U.S. counties from 2/24/2020 to 3/25/2020 and found that a rise in the local infection rate from 0% to 0.003% is associated with a 2.31% reduction in mobility. These smart-device based mobility statistics are also utilized to understand the effect of control measures in China [7] and to estimate the number of the Covid-19 infections in New York [8].

As for the Japanese case, Yabe et al. [9] employed 200,000 anonymized mobile phone users in Tokyo and concluded that by April 15^th^ (one week into the state of emergency), human mobility behavior decreased by approximately 50%. Similarly, Arimura et al. [10] analyzed the case of Sapporo City, Japan to understand the effect of the emergency declarations from the local and national government.

Our approach also utilizes mobility data (hourly and 500m grid scale populations across Japan) considering its powerful ability to capture the actual rate at which people stayed at home during the pandemic crisis. The goal of this study is to perform two major exploratory data analyses. First, we pick up the question, besides stay-at-home rates, what types of mobility measures are more correlated with infection risk? For this purpose, recent business census data at a 500m grid scale is combined with mobility statistics and the effective reproduction number (how many people are infected by one infected person, denoted by R(T)). The second focus question is, to what degree do we need to restrict our (daily) travel to reduce the pandemic crisis?

Compared to previous research with similar aims [2][3][7][8], the data set is constructed at a very detailed spatial scale and substantial effort is put into data processing to provide evidence that reinforces/complements the results in the literature. The effective reproduction number, estimated in this study is also helpful for interpreting the results because it is a direct indicator of an increasing or decreasing trend in infections. Understanding the relationship between mobility levels and the effective reproduction number can help policymakers make informed policy choices to reduce local infections.

### Data set and approach

In this study, three different types of statistics are utilized: the number of infected people [11], mobility statistics [12], and business census data from Japan’s Ministry of Internal Affairs and Communications (MIC) [13]. The number of infected people is recorded on the date that the infections are confirmed by Japan’s Ministry of Health, Labor and Welfare (MHLW). Here, we focused on the eight prefectures where the number of infections exceed500 by May 31, 2020. We then prepared the associated data sets for these eight prefectures.

Fig 1 illustrates the time series for the number of infected people in the eight target prefectures. Tokyo had the most infected people, and the second largest city, Osaka, follows. Explosions of infections can be seen from the end of March. It is assumed that people reduced their restriction levels during the holidays before this large wave came. April 7th is the day the emergency statement was issued by the Japanese Government, after which the first wave of the pandemic gradually abated.

**Fig 1.**
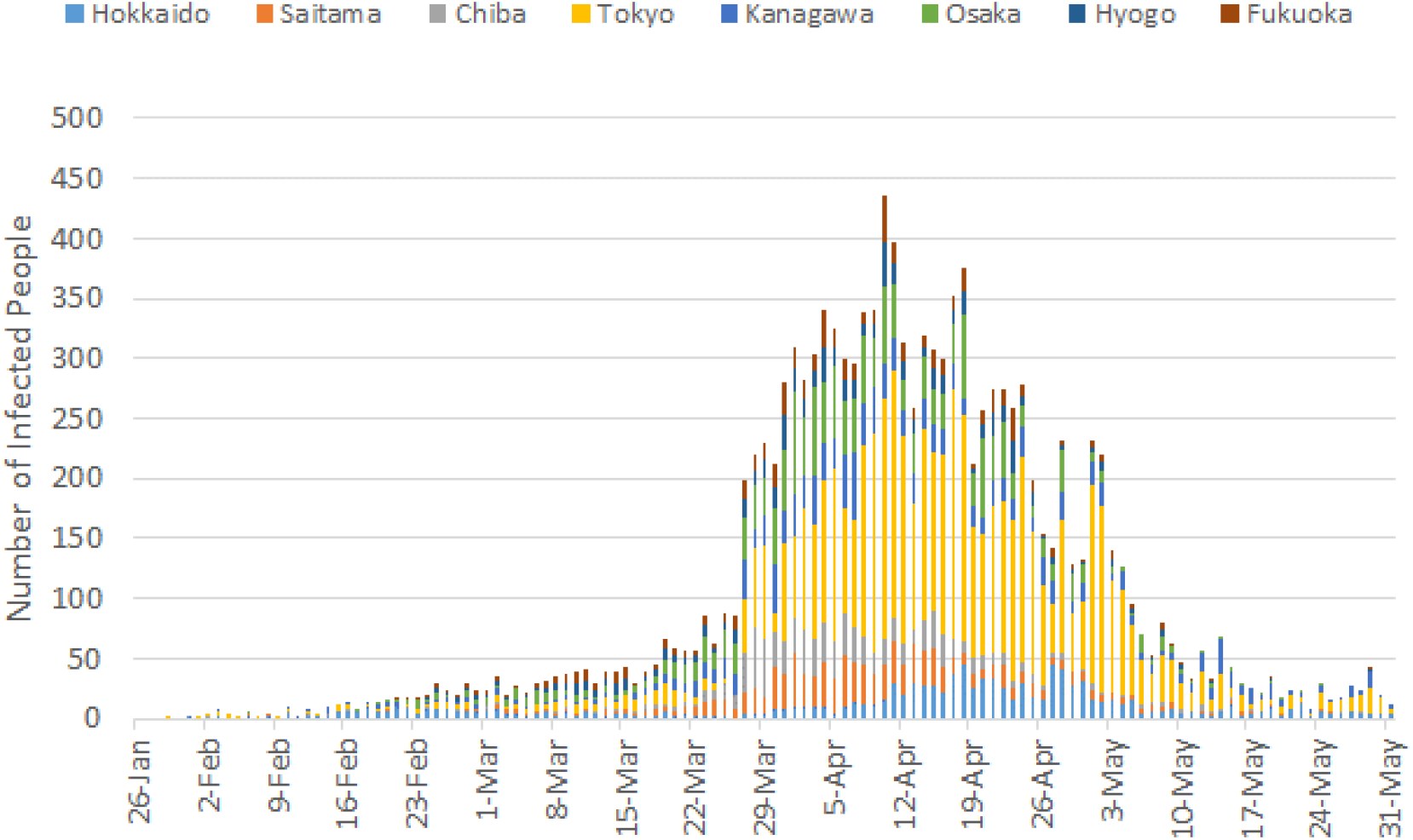
**Number of Infected People [11].**

The effective reproduction number (or instantaneous effective reproduction number due to time dependent characteristics) can be calculated from this data and the serial interval distribution (time between successive infections from one person to another). Basically, the infection process is normally regarded as a stochastic phenomenon due to the difference in infectivity among people and observation errors, and a certain type of probability distribution needs to be assumed. To estimate the effective reproduction number, we employed the model provided by Cori et al. [14], which assumes a gamma distribution for the number, the researchers validated the model by checking the consistency of the estimates for five historical outbreaks. We adopted Nishiura et al. [15] for the serial interval distribution: 4.8 days for the mean and 2.3 days for the standard deviation, which are relatively consistent with the report by Kramer et al. [7], which estimated 4.8 days for the mean and 3.3 days for the standard deviation.

NTT docomo is one of the largest carriers in Japan, and it holds 37.4 % of all mobile phone contracts in Japan [16]. Populations of people ages 15 to 79 are counted with the following rule for target “hourly duration” (i.e., one day is divided into 24 time slots such as 15:00-16:00.): If a person stays within the grid for 15 minutes during a target hour, then 1/4 is added to the population. The rule is applied for every 15 minutes of stay. Based on the duration of each person’s stay in a grid, either 1/4, 1/2, 3/4, or 1 is added to the population. The mobility data are not raw data, but are magnified based on the carrier’s, NTT docomo, share in each region. People below age 15 and over 80 as well as people that do not have a smartphone are not counted. Therefore, the estimated population tends to be lower than the actual population, but the majority of the population is explained by this data set.

Fig 2 depicts the mobility statistics in Tokyo for a single hour, 15:00-16:00, on two different days: On March 13 (Friday), a relatively dense population is observed, while in the same time period on Apr 23 (Friday), the population was reduced in central Tokyo. In Fig 2-1, the total number of people observed in the grids during the time period in March is 12,943,780 people, while a 2015 national census survey, the most recent data available, indicates a daytime population in Tokyo of 15,920,405 (Tokyo metropolitan government, 2018 [17]). In Fig 2-2, 12,128,864 people were counted in April. Reductions are seen in inflows from other prefectures and abroad as well as in inflows from residential areas to the center of the prefecture.

**Fig 2-1.**
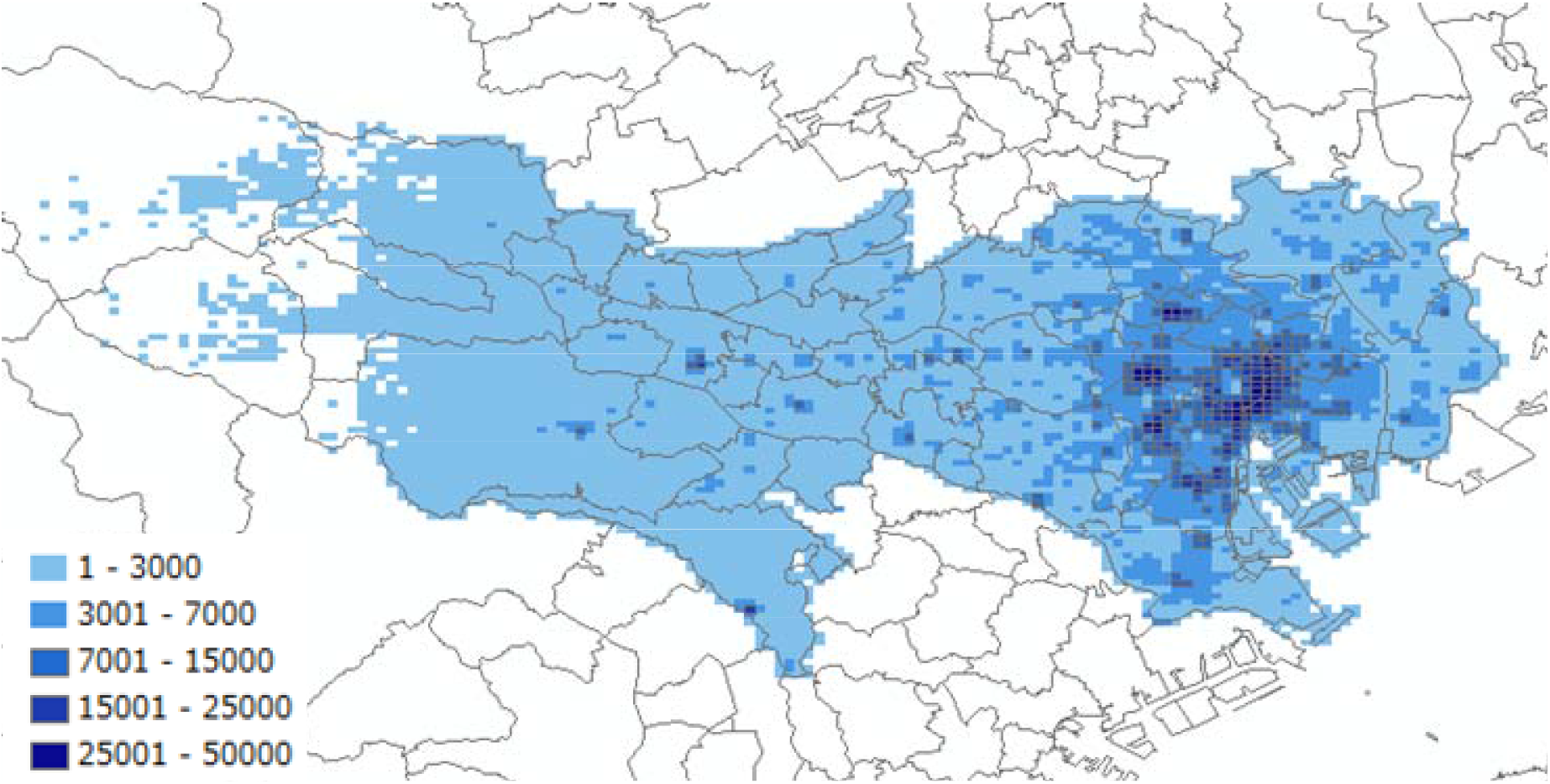
Mobility statistics in Tokyo at 15:00 (Mar 13)

**Fig 2-2.**
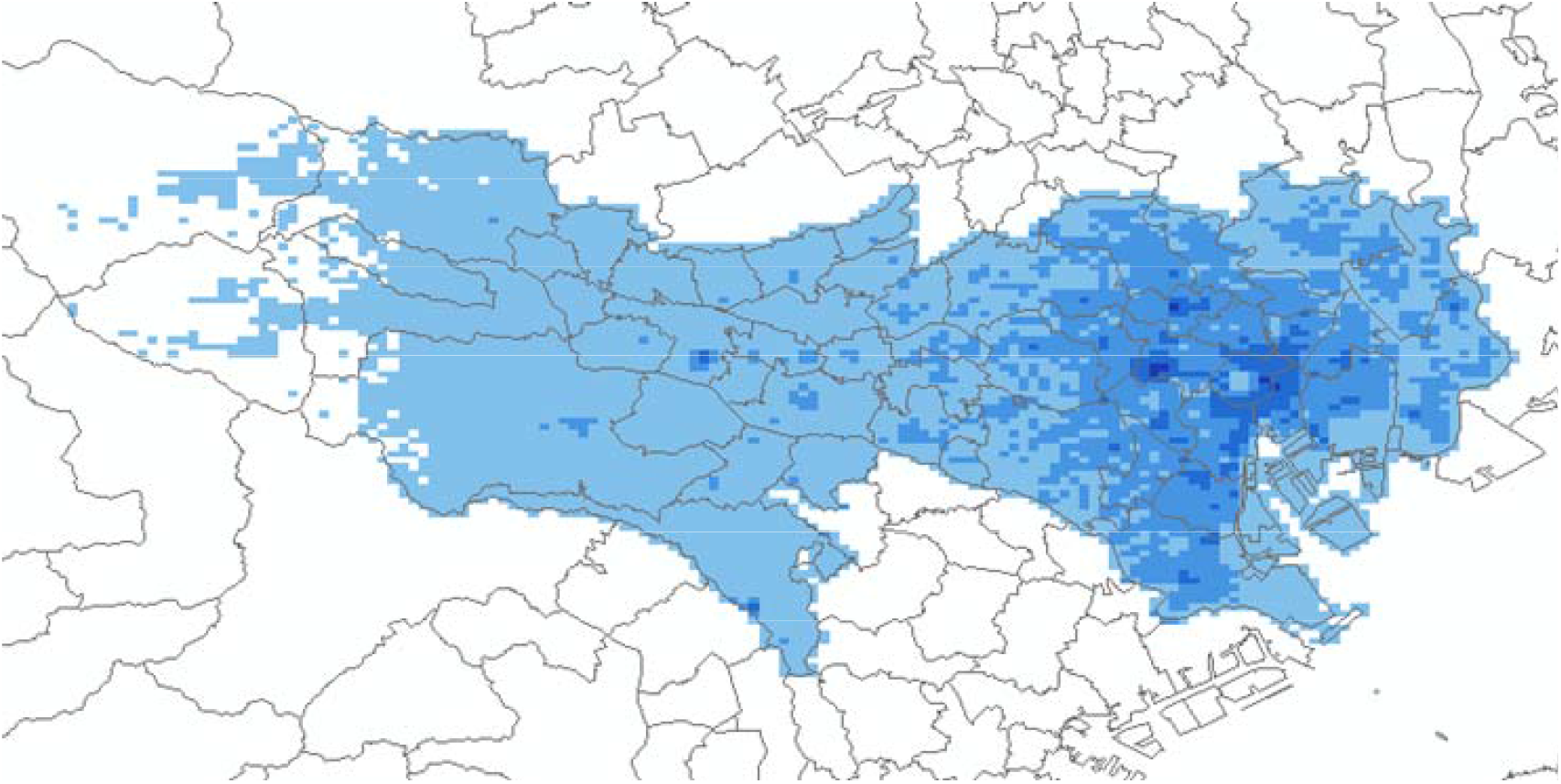
Mobility statistics in Tokyo at 15:00 (Apr 23)

The last statistic introduced is the 2016 economic census for businesses conducted by the MIC [13], which is the most recent data set available. The statistics include the number of employees in over 100 business sectors, and it is aggregated in a 500m grid scale. By using the mobility statistics and business census, we employ the following criterion as a measure of potential contacts in the business and commercial districts. This criterion should have a strong (negative) correlation to the level of stay home activity. The measure of (daily) potential contacts (PC) in the business and commercial districts is defined as the sum of the weighted average of the hourly population as follows:

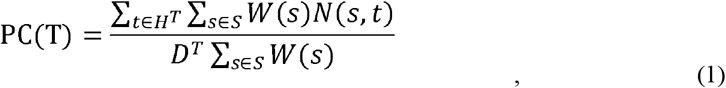

where *W*(*s*) is a variable for the weight at grid *s*, and *N*(*s, t*) is the mobility statistics at grid *s* and time *t*. In the analysis, we investigate two cases: *W*(*s*) = *TEmp*(*s*) as the total employees in all business sectors and *W*(*s*) = *HEmp*(*s*) as the total employees in all hospitality sectors (wholesale and retail, hotel and restaurant, living related and personal services, amusement, education, and medical and healthcare sectors). In either case, if people are crowded in business and commercial areas, where the number of employees is large, the value of the measure increases. This measure cannot capture the actual contacts (e.g., distance, meeting duration, mask use, and other countermeasures), but it is expected to explain the potential contacts considering that that literature has shown that a decrease in trips leads to a statistical decrease in infections.

The daily effective reproduction number R(T) is estimated over a weekly sliding window before T (i.e., R(T) represents the number from day T-6 to T). PC(T) is also estimated as a weekly average (from T-6 to T), but a 5 days lag is used to comparing R(T) (i.e., the incubation time is approximately 5 days, based on previous studies [19]). For simplicity, we denote the weekly average of PC(T) as 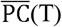. The period for estimating R(T) and 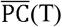 can be changed, but a good correlation is seen so far between R(T) and 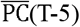 in the later analysis.

Based on the estimates of 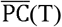 and R(T), the mobility restriction levels required to reduce the pandemic can be calculated. That is, the threshold value of 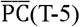that can achieve R(T) ≤1, is regarded as a minimum activity level. (If the 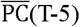 is smaller than the threshold, all the observed R(T) are smaller than or equal to 1.) If the threshold values of 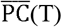 and *N*(*s, t*) are set as 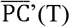 and *N*’(*s, t*), respectively, and the values of 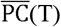 and *N*(*s, t*) at the normal period are set as 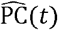 and 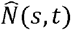, respectively, then the relative ratio of PC’(*t*) to 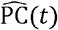 is given as:

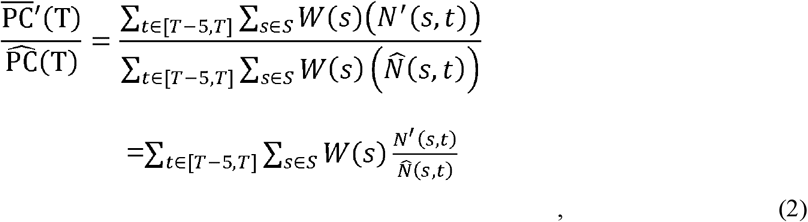

where N’(*s*,t) is the required population at grid *s* at time *t*, which is one of the solutions to achieve the threshold value. From Equation (2), we can understand that one of the solutions to achieve the target 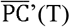 is to set the relative population 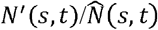 equals to 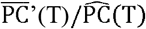 for all *s*ϵ S and *t*ϵ T.

## Results and discussion

We adopt the Pearson correlation coefficients to investigate the relationships between 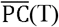 and R(T). To use the Pearson correlation coefficients, both variables must satisfy normality or linearity conditions. Therefore, we have applied the Kolmogorov-Smirnov test^1^ to 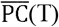 and R(T), respectively. As a result, we set the target period to the days from Apr 1^st^ to May 6^th^ (from about one week before the emergency declaration in the severely affected region to the end of the long holiday), which is regarded as the duration of the first severe wave. Unfortunately, the 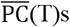 for two prefectures (Chiba and Saitama) did not pass the normality test (reject the null hypothesis on normality with 5 % significance level), but the other data sets are regarded as a normal distribution. The results of Chiba and Saitama are similarly analyzed as those in other prefectures in the later analysis for reference purpose only.

Fig 3 describes the Pearson correlation coefficients between R(T) and 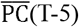 for two cases of employment (total and hospitality sector). From this figure, the correlation coefficients are generally better when employees in the hospitality sector are selected as the weight of 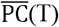. This indicates that infections tend to occur in the hospitality sector.

**Fig 3.**
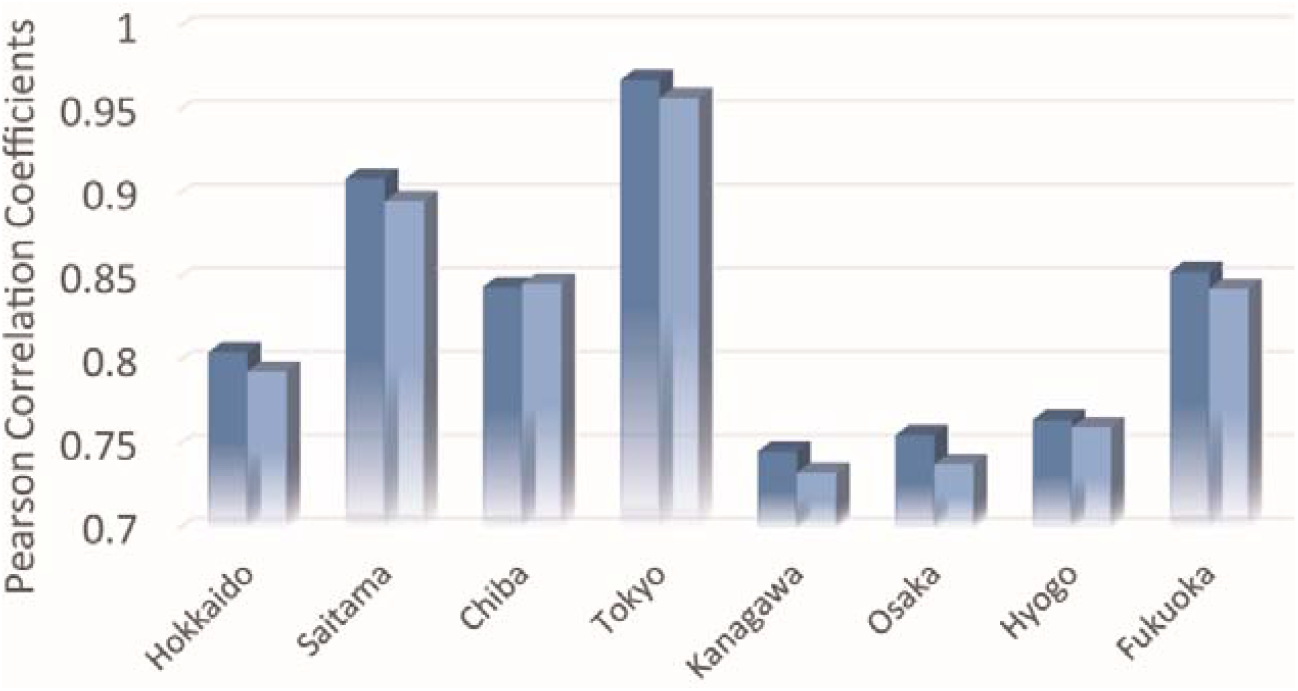
**Pearson correlation coefficients between effective reproduction numbers (R(T)) and the measure of potential contacts 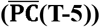 in the business and commercial district (applied to the daily data set from Apr 1 to May 6, 2020).**

Fig 4 plots the daily time series of 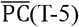 and R(T) in the eight prefectures. The number of infections was small during the first few weeks, and the R(T)s were not stable during this period. In the figure for Hokkaido, the target days after Apr1^st^ are highlighted for a reference. In many prefectures, the potential contacts decreased considerably in April and the beginning of May, but gradually started to return to pre-pandemic conditions at the end of May. The state of emergency declared by the central government ended on May 16^th^ in Hokkaido and on May 31^st^ in all the other prefectures, but people gradually restarted their activities, probably because the atmosphere of emergency was alleviated after many of the other prefectures ended emergency actions on May 6^th^.

**Figs 4-1-4-8. Fig 4.**
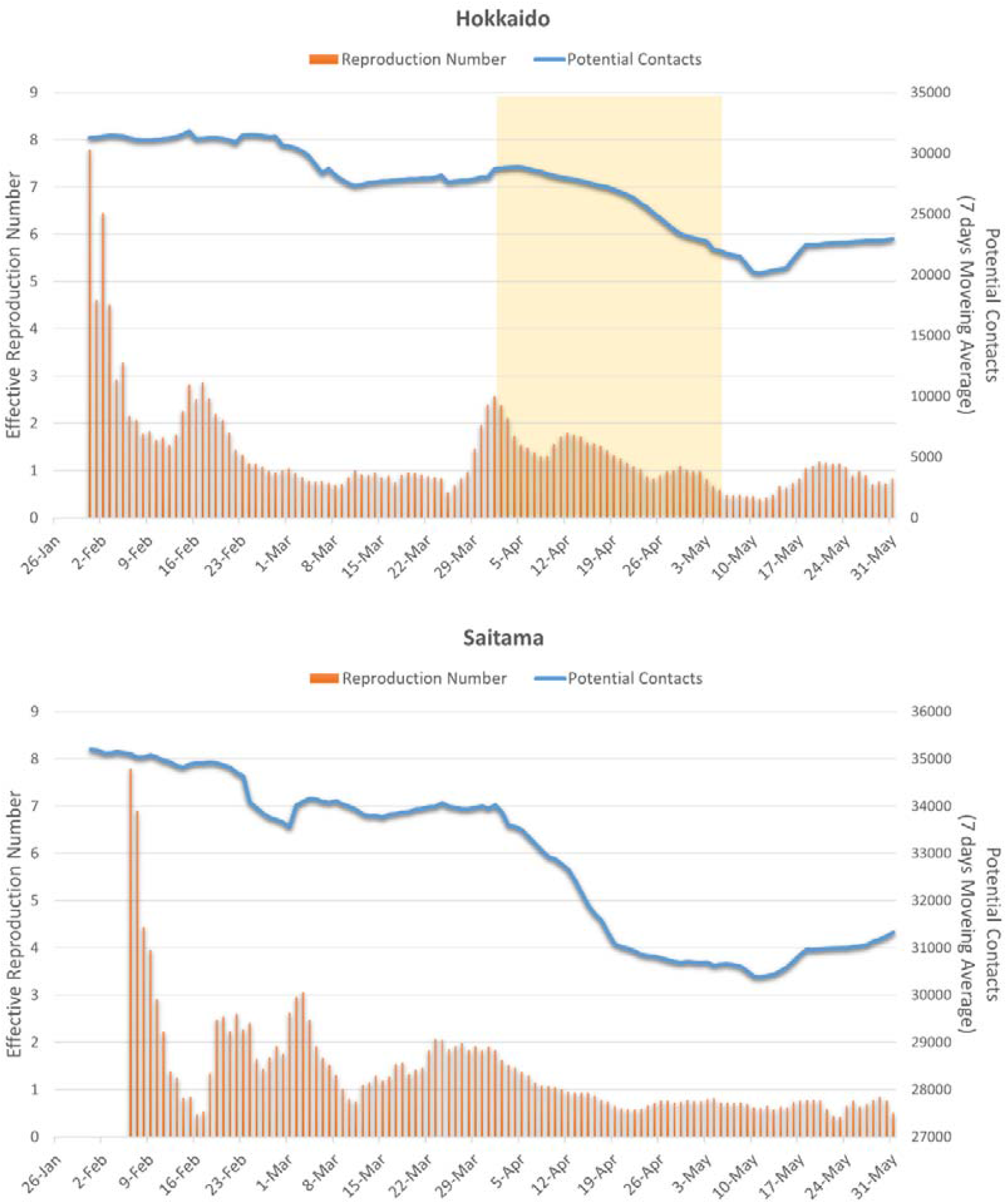
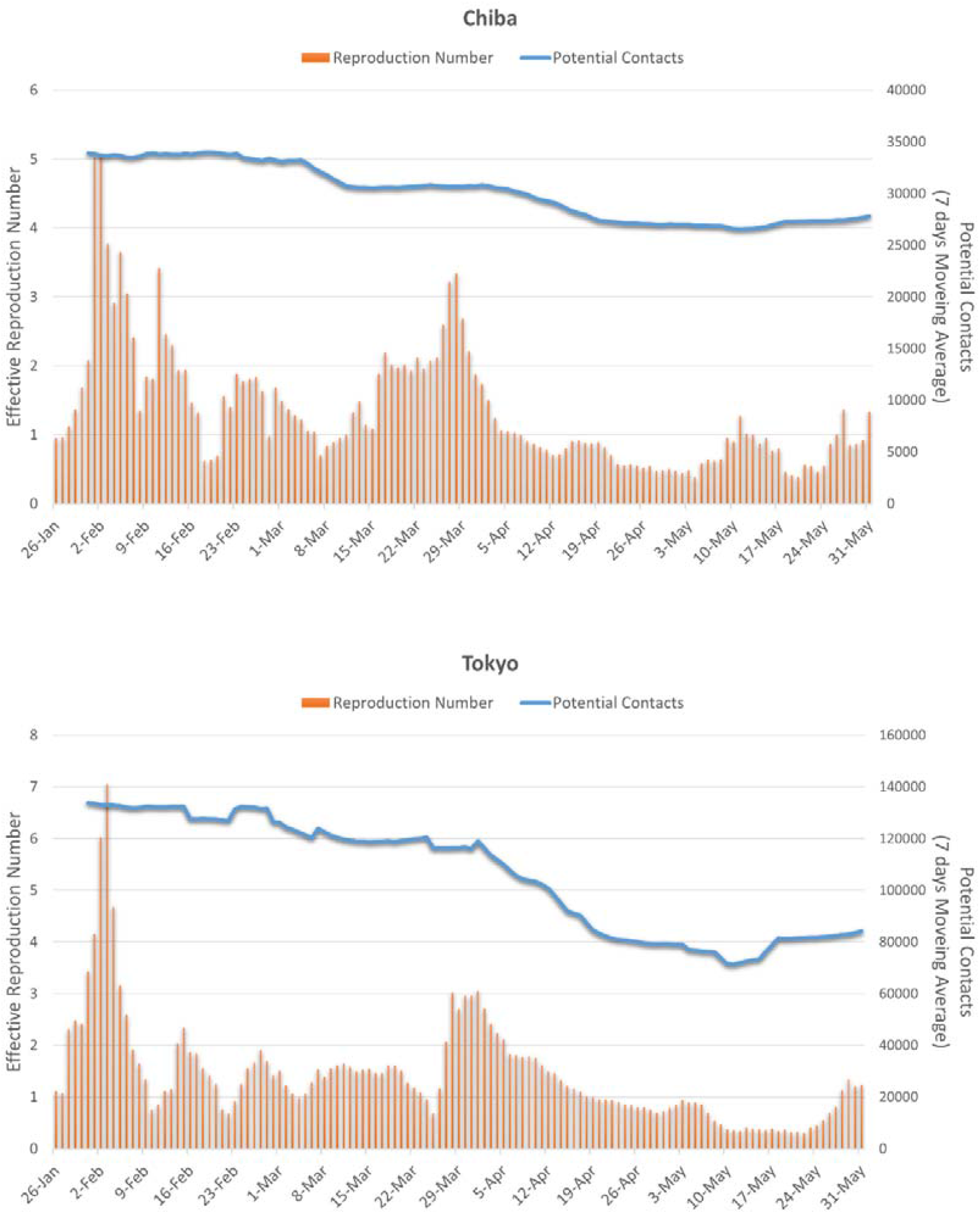
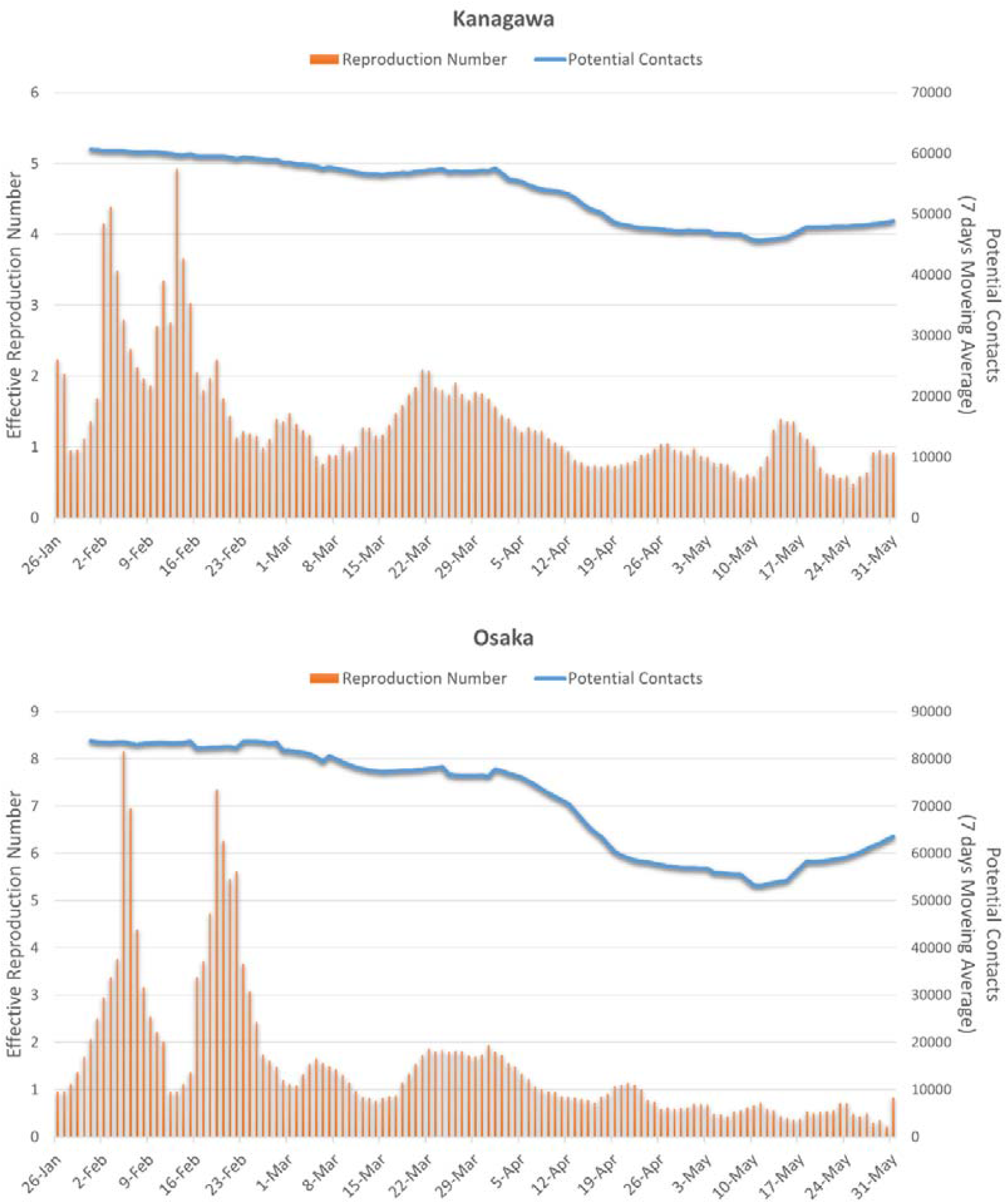
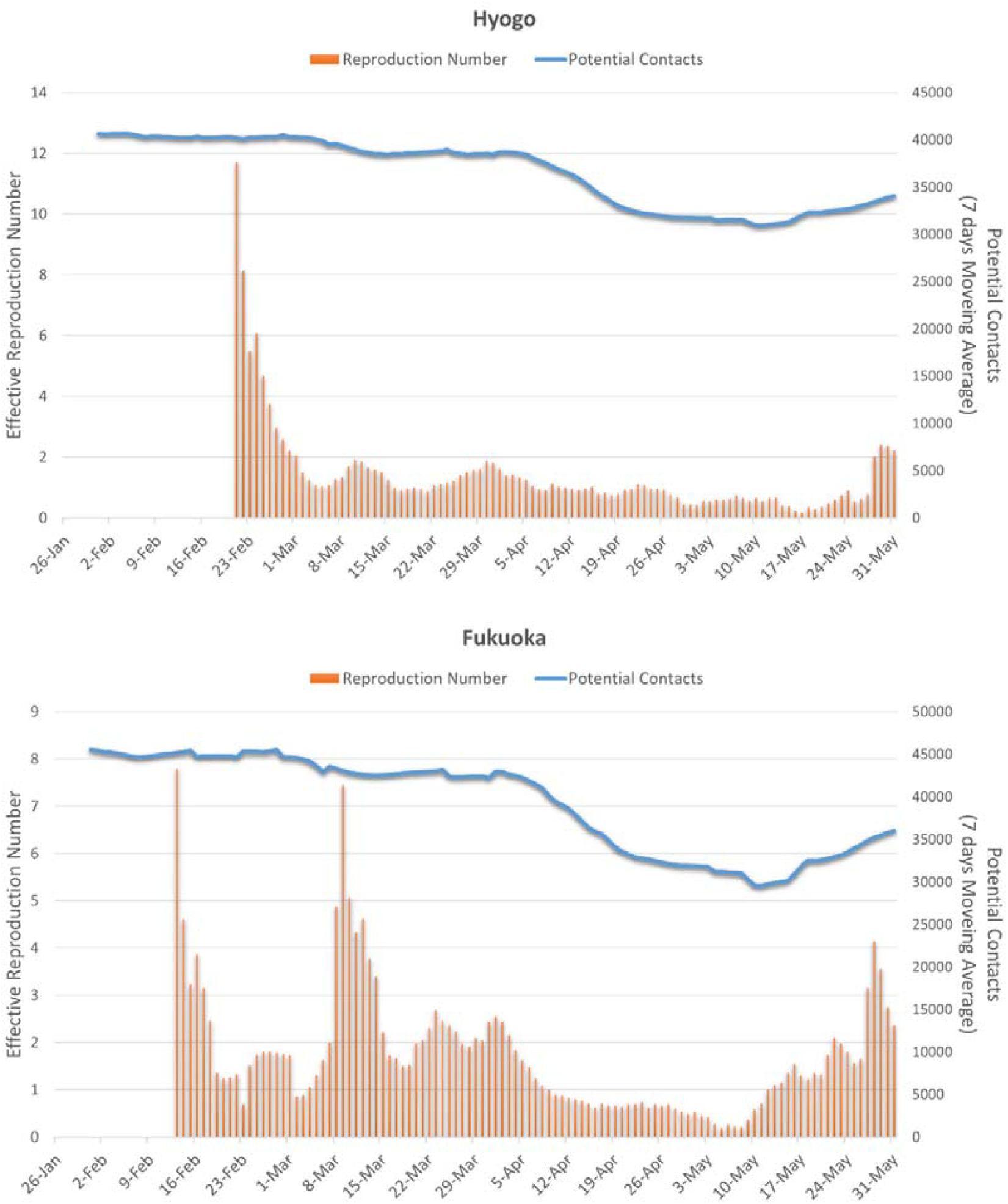
**Effective reproduction number (R (T)) and measure of potential contacts ((T-5)) in the eight prefectures (5 days lag for potential contacts is assumed, e.g., (T-5) on March 27^sh^ is plotted on April 1^st^)**

Based on the estimates of 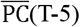 and R(T) described in Fig 4, the required mobility restriction levels can be calculated for reducing the pandemic from Equation (2). That is, the threshold value of 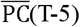 can achieve R(T) ≤1. Our study adopts the average 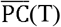 in February 2020 as the 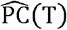 for Equation (2). Fig 5 shows the estimated population (mobility) restriction levels 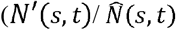 from *t*-6 to *t*) to ensure achievement of R(T) ≤1 in each prefecture. Because potential contacts are the sum of the weighted average of the population of employees in the hospitality sector, 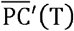 can be more easily achieved if the rate of people is reduced more in places where the hospitality sector is agglomerated.

**Fig 5.**
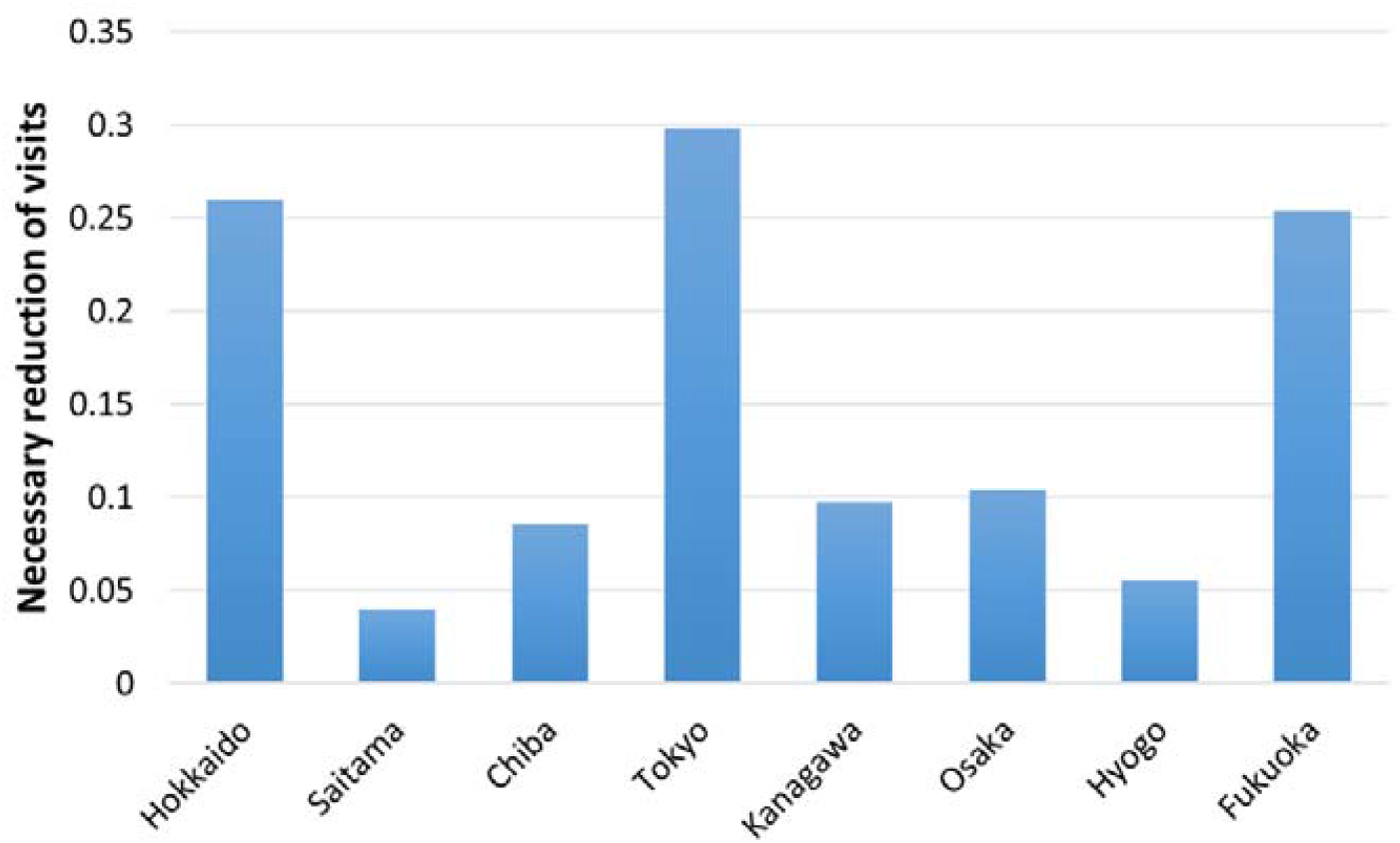
Necessary reduction level of visits to hospitality sectors to achieve R(t)<1 for “100% of days”.

From the figure, Tokyo requires the largest reduction (35%), followed by Osaka. This result can be naturally interpreted as these prefectures are the largest in Japan, and the population in the hospitality sector tends to be large. As shown in Fig 4, the scale of potential contacts in these prefectures is more than two times larger than the scale in the other prefectures. Hokkaido, Kanagawa, Hyogo, and Fukuoka also require high restriction levels on visits to the hospitality sector. Among these prefectures, Hyogo is a less populated prefecture, and the index of 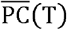 is low. Its population characteristics are generally reflected in the low value of R(T) in Hyogo, but a large restriction on visits to the hospitality sector is still required to guarantee R(t) ≤1. Another index, such as R(t) 1 with an 80% chance, may be appropriate to capture the relationships between 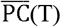 and an average low value of R(T). Saitama and Chiba may be classified into a third group, where the required restrictions are not so strict. However, these two sectors do not pass the normality test, and the relationship between mobility and the number of infections in these prefecture is not yet clear.

Based on the discussion above, in order to better understand the case of Hyogo, another index “R(T) ≤1 for 80% of days” is introduced to capture the relationships between 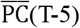 and R(t) in Fig 6. This provides a slightly different view from Fig 5. The required reduction level becomes much lower, especially in Kanagawa, Osaka, and Hyogo, where generally low R(T)s are observed. In these prefectures, a large number of infections were observed from an early stage, and the introduction of countermeasures (e.g., the number of people wearing masks and social distancing at local spots within a 500m grid) might have been preventative.

**Fig 6.**
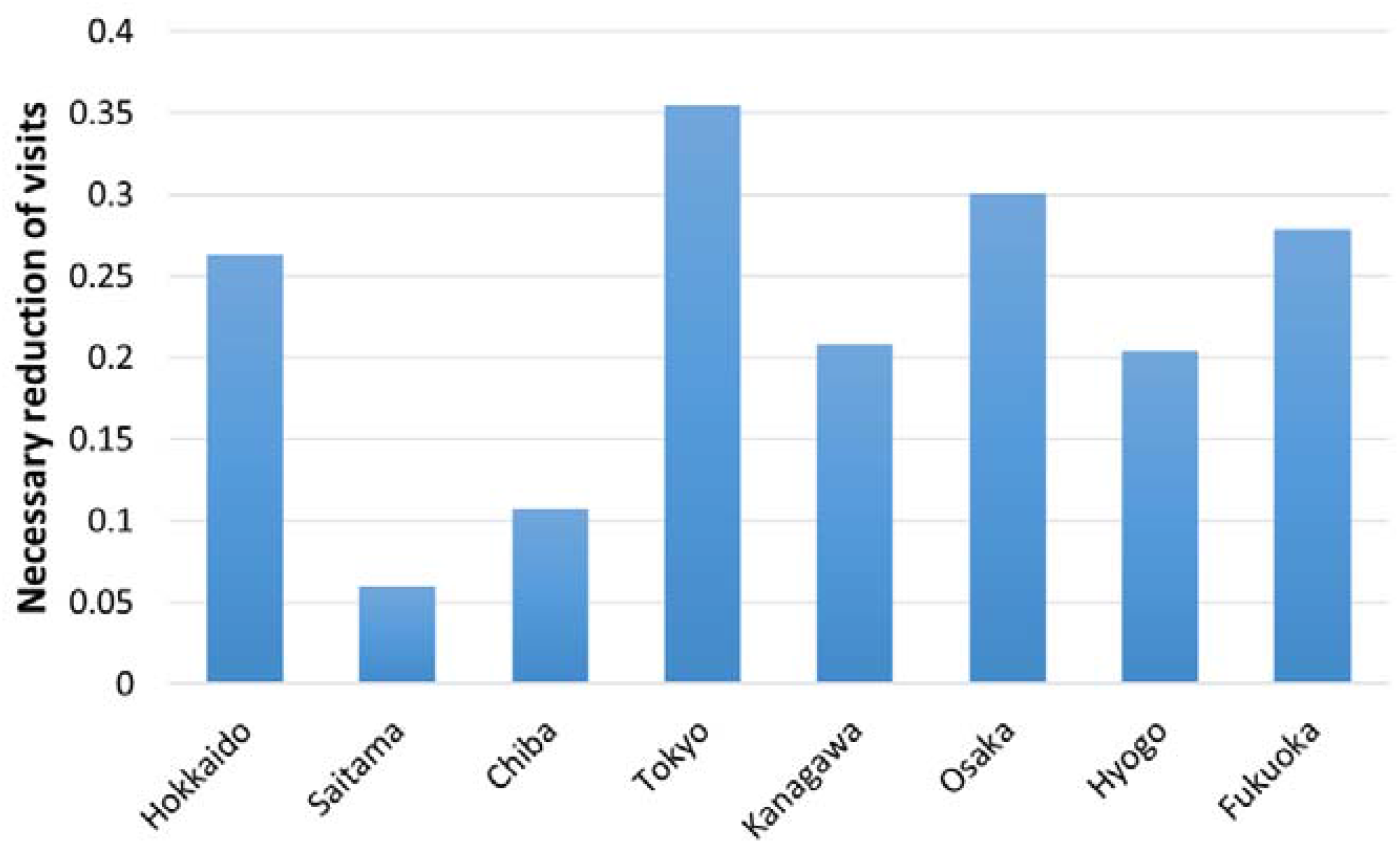
Necessary reduction level of visits to hospitality sectors to achieve R(t)<1 for “80% of days”.

On the other hand, Hokkaido, Tokyo, and Fukuoka still require a large reduction in the number of visits to the hospitality sector. Among these prefectures, Hokkaido and Fukuoka has a large agglomeration of hospitality sector businesses, especially restaurants and night spots for visitors, which possibly affected the infection rates.

The above result is based on a case in which an 80% reliability level is arbitrarily determined, but more discussion is needed to determine a reliability level that we can accept. A better discussion would be to investigate on the relationships between the mobility reduction levels at different reliability levels for R(T) ≤1 and to estimate the economic impacts of mobility reductions as well. However, this is beyond the scope of this study.

There are several studies that explain the change in total cases with a decrease in mobility. A quantitative comparison of these studies with our result is difficult because the data items and approach are different. For example, Glaser et al. (2020)[8] founds that 10% reduction in trips leads to a 0.27 log point drop in per capita Covid-19 prevalence. A 0.27 log point fall in Covid-19 represents five fewer cases per 1000 habitants, from a sample mean of 17 per 1000. Roughly speaking, 29.4% (=5/17) of cases are reduced by 10% reduction in trips. We interpret that mobility reduction leads to the effective reduction of new incidences. Their approach is favorable because the number of essential workers, which cannot avoid visiting areas with large numbers of infections, are considered an instrumental variable. Considering that our results indicate the reduction level needs to be around 20.3-35.4% to reduce new incidences, we cannot clearly say that lower percentage of mobility reduction would have a large potential to reduce new infections. We agree, at least, that the effects of mobility reduction have a statistical relationship with the number of infections, but varies depending on population density, especially in the hospitality sector.

Another necessary discussion point lies in the difference between the first wave treated in this study and the second or higher round waves. Considering that the countermeasures have advanced and the temperature has changed, further analysis is needed to know how much restrictions on mobility achieve R(T) ≤1. Continuous monitoring is necessary to understand when we will establish a new life with COVID-19.

## Conclusions

This study utilized mobility statistics and a business frame census, analyzed on a fine spatial scale, to capture the effective reproduction number of COVID-19, which is an important indicator in epidemiology. The weighted average of population density is estimated as a measure of congestion by using employees in the hospitality sector/total business sector. The study examines the correlation between these measures and the effective reproduction number in eight Japanese prefectures, where the incidence is large. One of the major conclusions in this study is that the measure of potential contacts in the hospitality sector (weighted average of population in the hospitality sector) has a fair correlation with the effective reproduction number.

From this measure, the necessary population reduction level to hold back the pandemic can be derived. Our analysis indicated 0.20 (Hyogo)-0.35 (Tokyo) reductions are required to achieve R(T) 1 for all days, depending on the conditions of the prefectures, but 0.06 (Hyogo)-0.30 (Tokyo) are enough to achieve R(T) 1 for 80% of days. Because of the regional variety in values, and the high sensitivity to the required reliability to achieve R(T) 1, these relationships should be carefully checked in each prefecture to determine mobility restriction policies. An analysis of the relationships between mobility reduction and economic impacts would also assist in this kind of policy making.

However, there are many limitations in the current study. As discussed, there are more explanatory variables with regard to natural and social conditions, which should be included in the analysis. Our analysis focused only on population density and business sector locations, but the attributes of the population are unknown. For example, age and job type largely affect the type of activity in the visited area. In addition to mobility and personal attributes, the number of incidences should be analyzed on a more detailed spatial scale. The current study uses the total number on a prefecture scale, which has a less significant relationship with mobility information in specific areas, especially when number of incidences is small. Delays on reported infections also affect the results in our study, and necessary modifications are required.

Moreover, for additional future studies on Japanese conditions, a comparative study between the first and second or higher round waves will be important to identify the progress of countermeasures and the effects of temperatures. A similar analysis in other countries would also help to understand what level of mobility restrictions and local countermeasures would contribute to a low infection risk.

## Data Availability

The mobility data underlying the results presented in the study are available as sales
products from NTT docomo InstightMarketing Inc. Other data sets are publicly available.

## Acknowledgements

We are grateful to NTT docomo InsightMarketing Inc. for providing mobility data sets in this study.

## Footnotes

1 This test investigates the hypothesis regarding whether the empirical distribution (observations) and theoretical distribution are statistically identical. A detailed explanation is given, for example, by Massey (1951)[18].

